# COVID-MATCH65 – A prospectively derived clinical decision rule for severe acute respiratory syndrome coronavirus 2

**DOI:** 10.1101/2020.06.30.20143818

**Authors:** Jason A Trubiano, Sara Vogrin, Olivia C Smibert, Nada Marhoon, Adrian A Alexander, Kyra Y L Chua, Fiona L James, Nicholas RL Jones, Sam E Grigg, Cecilia LH Xu, Nasreen Moini, Sam R Stanley, Michael T Birrell, Morgan T Rose, Claire L Gordon, Jason C Kwong, Natasha E Holmes

**Affiliations:** Department of Infectious Diseases, Austin Health, Heidelberg, Australia; Department of Medicine (Austin Health), University of Melbourne, Heidelberg, Australia; Department of Infectious Diseases and The National Centre for Infections in Cancer, Peter MacCallum Cancer Centre, Parkville, Australia; Department of General Medicine, Austin Health, Heidelberg, Australia; Department of Medicine (St Vincent’s Hospital), University of Melbourne, Fitzroy, Australia; Data Analytics Research and Evaluation (DARE) Centre, Austin Health and University of Melbourne, Heidelberg, Australia; Electronic Medical Record and Information and Communications Technology Services, Austin Health, Heidelberg, Australia; Department of Microbiology and Immunology, University of Melbourne, Peter Doherty Institute for Infection and Immunity, Melbourne, Australia

**Keywords:** SARS-CoV-2, diagnosis, PCR, risk

## Abstract

Due to the ongoing COVID-19 pandemic and increased pressure on testing resources, understanding the clinical and epidemiological features closely associated with severe acute respiratory syndrome coronavirus 2 (SARS-CoV-2) is vital at point of care to enable risk stratification. We demonstrate that an internally derived and validated clinical decision rule, COVID-MATCH65, has a high sensitivity (92.6%) and NPV (99.5%) for SARS-CoV-2 and could be used to aid COVID-19 risk-assessment and resource allocation for SARS-CoV-2 diagnostics.

## Background

The COVID-19 pandemic caused by severe acute respiratory syndrome coronavirus 2 (SARS-CoV-2) was first reported in China and has now infected over 9 million people globally (1). A range of clinical symptoms and syndromes have been reported in confirmed COVID-19 (2-4). However, there have been limited prospective reports of the clinical and epidemiological predictors of COVID-19 infection (5). We report on the clinical and epidemiological predictors of COVID-19 from a uniquely derived prospective database and present a point-of-care COVID-19 clinical decision tool.

## Methods

A COVID-19 rapid assessment screening clinic was established at Austin Health on 11 March 2020 with prospective electronic medical record (EMR; **eMethods**) data of patients presenting to the clinic systematically collected by medical staff from 11 March to 22 April 2020. Patients were predominantly adults - children over 6 months were seen at clinician discretion. Modifications to the EMR were made during the study period to align with the Victorian Department of Health and Human Services (DHHS) testing criteria (6) (**eMethods**). Only those patients that met the DHHS criteria for SARS-CoV-2 testing had nasopharyngeal swab collected for SARS-Cov-2 nucleic acid detection by polymerase chain reaction (PCR). Patients with swabs that had SARS-CoV-2 nucleic acid detected were termed “COVID-19 test positive”; those with swabs where SARS-CoV-2 nucleic acid was not detected were termed “COVID-19 test negative”. This study was approved by the Austin Health Human Research and Ethics Committee.

### Derivation and Internal Validation Cohort

Clinical data from the data collection tool (baseline demographics, clinical symptoms, clinical observations) and COVID-19 testing results were extracted from Austin Health EMR platform (Cerner®) by the Data Analytics Research and Evaluation (DARE) Centre (Austin Health/University of Melbourne).

### Statistical analysis

All results are presented according to TRIPOD guidelines(7). Categorical variables are presented as frequency (percentage) and continuous variables as median (interquartile range [IQR]). Fisher’s exact test or rank sum test were used to compare characteristics between tested and not tested patients. To determine the predictors of a positive COVID-19 test, a multivariable logistic regression with backward stepwise procedure was used, eliminating variables with p>0.10 and re-inclusion of variables with p<0.05. Bootstrapping was used for internal validation. Further details on variable selection, model development and performance, internal validation and score derivation are outlined in **eMethods**.

## Results

### Study population and setting

During the study period 4359 assessments were performed in 4226 patients (**eTable 1**). For those with multiple presentations (n=118) only their first testing date was used (for patients that were not tested, their first assessment was taken). Median (IQR) number of daily assessments was 96 (71, 134) with an average of 51% of patients being tested each day (**eFigure1**).

### COVID-19 testing

Testing was performed on 2976 patients (70%). The characteristics of those with suspected COVID-19, stratified by testing performed status, is outlined in **eTable 2**. The most frequently reported symptoms in both groups were any fever (reported or documented), cough, sore throat and coryza as outlined in **eTable 2**.

### COVID-19 test positivity

Of the 2976 patients that were tested, 41 were excluded from the analysis due to pending results (n=38) or indeterminate results (n=3). The prevalence of a positive COVID-19 test in the final cohort was 3.7% (108/2935). Characteristics of those patients with a positive COVID-19 test are shown in **Table 1**.

**Table 1.**
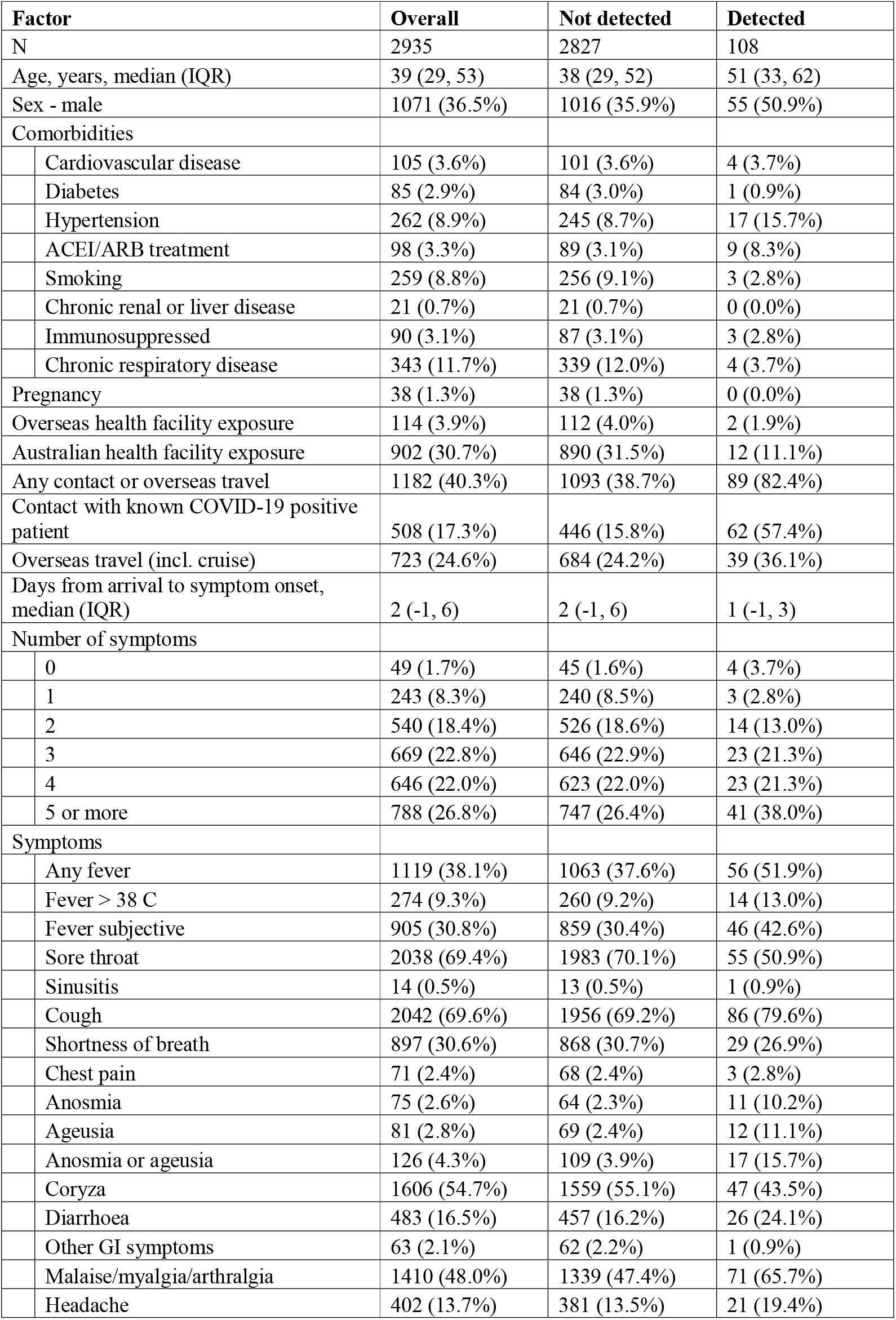
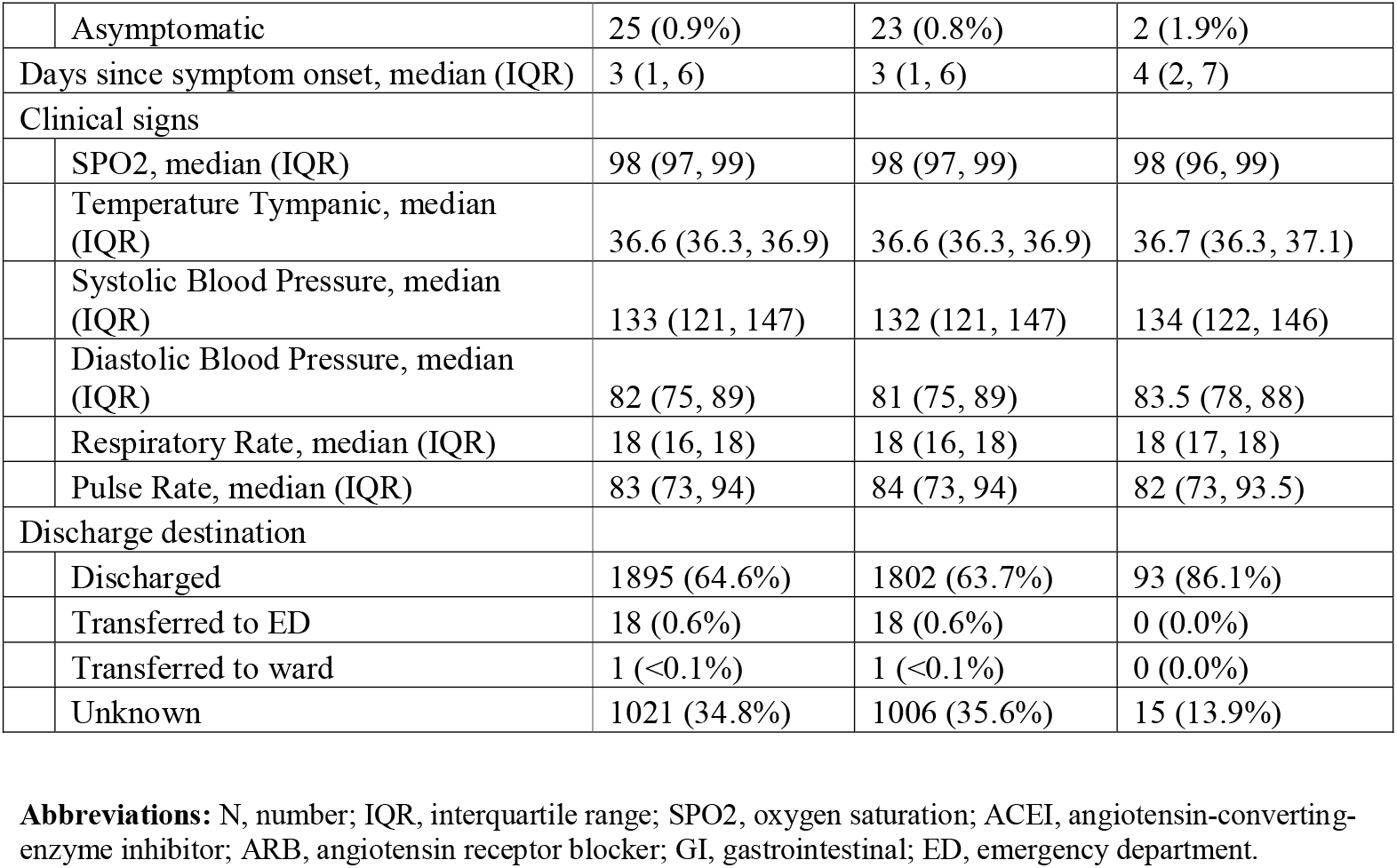
Characteristics of patients who underwent testing for COVID-19

### Demographic, epidemiological and clinical factors associated with a positive COVID-19 test

The characteristics associated with a positive COVID-19 test in univariate and multivariable analysis are shown in **Table 2**. The seven features associated with a COVID-19 test on multivariable analysis were summarized in the mnemonic COVID-MATCH65 (**Figure 1**). The model showed good discrimination (AUC = 0.843, Hosmer-Lemeshow chi^2^=4.96, p=0.762) and calibration (calibration slope = 1.00, Brier score = 0.03, product-moment correlation between observed and predicted probability = 0.35). Internal validation showed minimal mean optimism of 0.007 with internally validated AUC of 0.836 **(eFigure 2 & 3)**. The resulting score ranges from -1 to 6.5 points with score ≤ 1 representing low risk of a positive test (<1%) and scores above 4 having beyond 20% probability of a positive test (**Figure 1**).

**Table 2:** Univariate & multivariable analysis of features associated with a positive COVID-19 test (SARS-CoV-2 nucleic acid Detected)

| Variables considered for inclusion | Overall | COVID-19 positive test | Univariate analysis |  | Multivariable analysis |  |  |  | Points |
| --- | --- | --- | --- | --- | --- | --- | --- | --- | --- |
|  |  |  | OR (95% CI) | p-value | OR (95% CI) | Beta coefficient (95% CI) | p-value | Presence in 1000 bootstrap replications, %* |  |
| Age 65+ | 254 (8.7%) | 19 (17.6%) | 2.35 (1.41, 3.93) | 0.001 | 2.80 (1.56, 5.04) | 1.03 (0.45, 1.62) | 0.001 | 75 | 1 |
| Male sex | 1071 (36.5%) | 55 (50.9%) | 1.85 (1.26, 2.72) | 0.002 |  |  |  | 50 |  |
| Hypertension | 262 (8.9%) | 17 (15.7%) | 1.97 (1.15, 3.36) | 0.013 |  |  |  | 50 |  |
| Contact with known COVID-19 positive patient or overseas travel | 1182 (40.3%) | 89 (82.4%) | 7.43 (4.50, 12.27) | <0.001 | 14.24 (7.92, 25.63) | 2.66 (2.07, 3.24) | <0.001 | 100 | 2.5 |
| Any fever (documented or reported) | 1119 (38.1%) | 56 (51.9%) | 1.79 (1.22, 2.63) | 0.003 | 1.59 (1.03, 2.43) | 0.46 (0.03, 0.89) | 0.035 | 71 | 0.5 |
| Coryza or sore throat | 2455 (83.6%) | 73 (67.6%) | 0.39 (0.26, 0.59) | <0.001 | 0.36 (0.23, 0.58) | -1.01 (-1.48, -0.55) | <0.001 | 99 | -1 |
| Cough | 2042 (69.6%) | 86 (79.6%) | 1.74 (1.08, 2.80) | 0.022 |  |  |  | 52 |  |
| Shortness of breath* | 897 (30.6%) | 29 (26.9%) | 0.83 (0.54, 1.28) | 0.394 |  |  |  |  |  |
| Anosmia or ageusia | 126 (4.3%) | 17 (15.7%) | 4.66 (2.68, 8.09) | <0.001 | 13.67 (6.89, 27.13) | 2.62 (1.93, 3.30) | <0.001 | 100 | 2.5 |
| Diarrhoea | 483 (16.5%) | 26 (24.1%) | 1.64 (1.05, 2.58) | 0.031 |  |  |  | 26 |  |
| Myalgia or Malaise | 1410 (48.0%) | 71 (65.7%) | 2.13 (1.42, 3.19) | <0.001 | 2.20 (1.41, 3.44) | 0.79 (0.45, 1.35) | 0.001 | 96 | 1 |
| Headache | 402 (13.7%) | 21 (19.4%) | 1.55 (0.95, 2.53) | 0.079 |  |  |  | 36 |  |
| SPO2 <97% | 473 (16.1%) | 36 (33.3%) | 2.73 (1.81, 4.13) | <0.001 | 2.46 (1.57, 3.87) | 0.90 (0.45, 1.35) | <0.001 | 93 | 1 |
| Temperature ≥37.5 C | 174 (5.9%) | 11 (10.2%) | 1.85 (0.97, 3.53) | 0.060 |  |  |  | 15 |  |
| Systolic blood pressure >140 mmHg* | 1082 (36.9%) | 43 (39.8%) | 1.14 (0.77, 1.69) | 0.518 |  |  |  |  |  |
| Diastolic blood pressure >80 mmHg | 1623 (55.3%) | 72 (66.7%) | 1.65 (1.10, 2.47) | 0.016 |  |  |  | 54 |  |
| Respiratory rate <16/min or >20/min* | 196 (6.7%) | 7 (6.5%) | 0.97 (0.44, 2.11) | 0.934 |  |  |  |  |  |
| Pulse rate <60/min or >100/min | 486 (16.6%) | 11 (10.2%) | 0.56 (0.30, 1.06) | 0.073 |  |  |  | 51 |  |

\*Not considered for inclusion due to p<0.200
**Abbreviations:** OR, odds ratio; CI, confidence interval; SPO2, oxygen saturation

**Figure 1:**
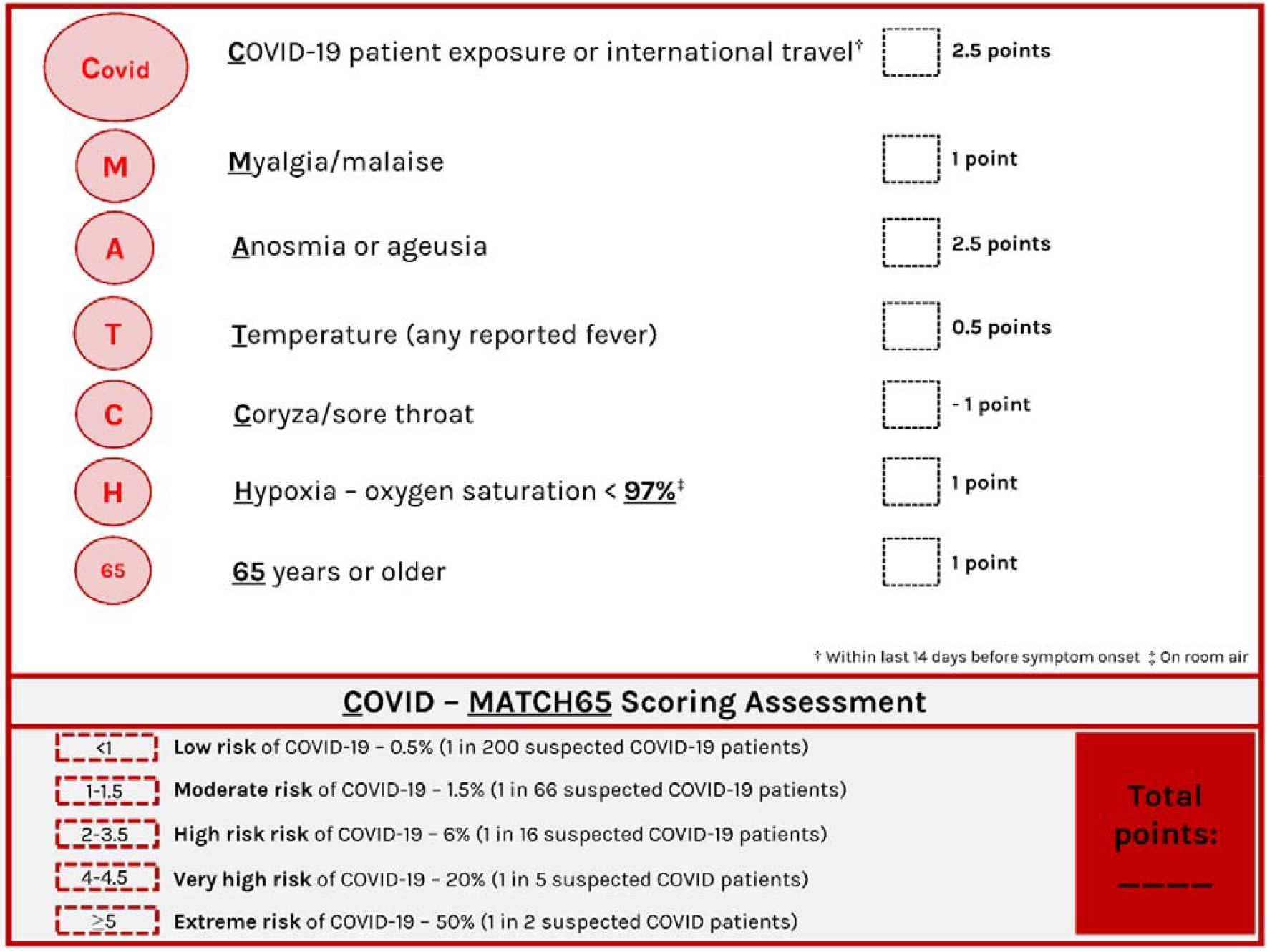
COVID-19 clinical decision rule – COVID-MATCH65

The positive and negative results for each COVID-MATCH65 score are outlined in **eTable 2**. A score of at least 1.5 was shown to have 92.6% (95% CI 85.9%, 96.7%) sensitivity, 51.3% (49.4, 53.1) specificity, 6.8% (5.5, 8.2) positive predictive value and 99.5% (98.9, 99.8) negative predictive value of identifying a patient who was COVID-19 test positive (**eTable 3**). COVID-MATCH65 also retains a high NPV with increasing prevalence of COVID-19 (30% prevalence) (**eTable 4**).

### Admission to hospital

A total of 15 COVID-19 positive patients (14%) were admitted to hospital. Median (IQR) COVID-MATCH65 score in admitted was 3.5 (2.5, 4.5) while in non-admitted it was 3 (2.5, 4). Score was not predictive of admission (OR 1.04, 95%CI: 0.70, 1.53, p=0.852). Variables predictive of admission were oxygen saturation (SpO2) < 97%, shortness of breath, male gender and not being exposed to confirmed case/international travel (**eTable 5**).

## Discussion

Whilst the clinical features of COVID-19 have been well reported, robust prospective from patients presenting for COVID-19 assessment that are both SARS-CoV-2 positive and negative on testing remains absence. Therefore, to date an accurate assessment of the clinical predictors associated with a positive SARS-CoV-2 test has been ill defined. Whilst fever has been the predominant presenting feature of confirmed COVID-19 cases from published inpatient populations(4), it was in fact observed less frequently (36.5%) in our outpatient cohort, potentially the result of earlier presentation (5 days[median] from symptom onset). Bajema *et al*.(5) reported fever in 68% in a retrospective cohort study (n=210) from the USA with similar incidence rate of COVID-19 positive tests to our cohort (5% USA vs. 4.7% AUS). Whilst in the earliest reports from confirmed cases in China the figures were 83-98%(2, 3). Whilst coryza and sore throat were frequently reported, the presence of either was in fact a negative predictor of COVID-19 infection. Anosmia or ageusia as seen in other emerging studies was a strong predictor of a positive COVID-19 test(8). Whilst contact and/or international travel was a predictor of COVID-19 infection in our model, as seen in US model from Challenger *et al*.(9), it may be less relevant in outbreak settings and during periods of travel bans, however these criteria alone are not required for a patient to be at high risk of COVID-19.

Our model has some limitations, including the single centre prospective data source, jurisdictional guided testing criteria, testing of symptomatic only patients and absence of external validation. However, only one small retrospective US cohort (n =49 COVID-19 positive /n= 98 COVID-19 negative)(9) and two non-peer reviewed publications from China have examined the role of clinical decision rules from large datasets - Meng *et al*. (n = 620 outpatients; 48.7% positive)(10) and Song *et al*.(11) (n = 304 inpatients; 24.0% positive), both limited by requirement for clinical and laboratory parameters. COVID-MATCH65 uses readily available clinical information without laboratory test results, with a score of 1.5 associated with high sensitivity and NPV, enabling application in the outpatient and potentially early inpatient setting. Further risk stratification can be made with the scoring tool (lowest risk [< 1 in 100] to extreme risk [1 in 1]), aiding diagnostic approaches in patients with suspected COVID-19. In a pandemic where diagnostic resources are limited in both low- and high-income settings,(12) risk stratification of those likely to have COVID-19 is urgently required and tools such as COVID-MATCH65 can aid the front-line clinician. We encourage readers to urgently employ and validate COVID-MATCH65 in their own datasets, as it is likely to aid clinicians at point-of-care especially via an open access web platform (http://COVID-MATCH65.austin.org.au).

## Data Availability

All data is available

## Author contributions

JAT, NEH, OS, JCK, KYC – Study design, manuscript preparation, manuscript review; SV – statistical analysis; AA, NM, MTB, SEG, CLX – data collection, manuscript review; MR, SS – data collection and database design; NJ, FLJ – manuscript review. All authors have read and approved the manuscript.

## Acknowledgements

Nil

